# Predicting non-response to ketamine for depression: a symptom-level analysis of real-world data

**DOI:** 10.1101/2023.07.03.23292094

**Authors:** Eric A. Miller, Houtan Totonchi Afshar, Jyoti Mishra, Dhakshin Ramanathan

## Abstract

**Background:** Ketamine helps some patients with treatment resistant depression (TRD), but reliable methods for predicting which patients will, or will not, respond to treatment are lacking.

**Methods:** This is a retrospective analysis of PHQ-9 item response data from 120 military veterans with TRD who received repeated doses of intravenous racemic ketamine or intranasal eskatamine in a real-world clinic. Regression models were fit to individual patients’ symptom trajectories and model parameters were analyzed to characterize how different symptoms responded to treatment. Logistic regression classifiers were used to predict treatment response using patients’ baseline depression symptoms alone. Finally, by parametrically adjusting the classifier decision thresholds, the full space of models was searched to identify the best models for predicting non-response with very high negative predictive value.

**Results:** Model slopes indicated progressive improvement on all nine symptoms, but the symptom of depressed mood improved faster than the symptom of low energy. The first principal component (PC) represented a data-driven measurement of overall treatment response, while the second PC divided the symptoms into affective and somatic subdomains. Logistic regression classifiers predicted response better than chance using baseline symptoms, but these models achieved only 60.2% predictive value. Using threshold tuning, we identified models that can predict non-response with a negative predictive value of 96.4%, while retaining a specificity of 22.1%, suggesting we could successfully identify 22% of individuals who would not respond purely based on baseline symptom scores.

**Conclusions:** We developed an approach for identifying a subset of patients with TRD who will likely not respond to ketamine. This could inform rational treatment recommendations to avoid additional treatment failures.

## Introduction

Despite effective treatments for depression, such as psychotherapy [1] and monoamine-based antidepressants [2], many patients do not find remission even after multiple trials of different treatments [3]. With a novel mechanism of action, ketamine offers hope for such patients with treatment resistant depression (TRD) [4]. However, our ability to predict which patients will respond to ketamine remains limited. Although studies have identified potential moderators of response, such as obesity [5,6], family history of alcohol use disorder [5,7], and concomitant benzodiazepine use [8], a recent large meta-analysis failed to detect any consistent patient-level predictors of response to ketamine [9]. In the absence of consistent predictors of response, it remains difficult to stratify and select the optimal treatment for a particular patient among a growing list of options for TRD, including electroconvulsive therapy (ECT) [10], transcranial magnetic stimulation (TMS) [11], magnetic seizure therapy (MST) [12], deep brain stimulation (DBS) [13], and psychedelic-assisted psychotherapy [14].

Prediction fundamentally rests on a choice of how to measure changes in the outcome of interest, in this case changes in depression. Most clinical studies utilize questionnaire sum scores to quantify depression before and after treatment, with response defined via a proportional drop in the sum score. Even for validated questionnaires, sum scores can mask individual differences or dynamics at the level of symptoms [15,16] or dimensional features of illness [17]. For example, different symptom clusters may have distinct etiological and physiological underpinnings, which may, in turn, respond differently to treatments [18,19]. Although many studies have reported on ketamine’s effects specifically on suicidality [20], we are aware of only a few studies that have compared how ketamine affects different depression symptoms or symptom clusters[21–23]. None of those studies have modeled trajectories of symptoms over time.

Our first goal, therefore, was to model how individual symptoms of depression change over time for patients undergoing repeated ketamine treatments. This is a secondary analysis of real-world clinical data from a population of military veterans, most of whom have numerous psychiatric comorbidities. Over two-thirds of the patients had a diagnosis of PTSD in addition to depression. As such, these data reflect how ketamine may be expected to perform for the treatment of TRD among complex patients, the very patients who could benefit most from novel and effective treatments. In smaller studies from the same population, our group has found that ketamine is effective for these patients, but with lower response rates than commonly reported in clinical trials [24,25]. This further highlights the need for reliable predictors of response or, perhaps more importantly, predictors of non-response to ketamine among complex patients.

As a secondary goal, building on our models of symptom trajectories, we used machine learning classifiers to predict whether patients would respond to ketamine using their baseline item by item PHQ-9 symptom scores. We also developed a model that can predict non-response, i.e. treatment failure, with very high confidence for a meaningful subset of the patients. These findings contribute to our understanding of how ketamine affects individual symptoms of depression for complex patients in a real-world clinical setting. Our method for identifying patients who are unlikely to respond to ketamine could prove useful, if replicated, for guiding treatment recommendations among a growing list of interventions targeting TRD.

## Methods

### Patients

Data were obtained from 120 patients who underwent serial ketamine induction treatments for depression at the San Diego Veterans’ Administration hospital between January 2020 and June 2022. 85 patients were male and 35 were female. Ages ranged from 26 to 75 (mean 45, standard deviation 12) years. Most patients (92%) carried a diagnosis of major depression, but comorbidity was very common. The most frequently co-occurring condition was PTSD in 73% of patients, with less frequent conditions including generalized anxiety disorder, bipolar disorder, ADHD, borderline personality disorder, and various substance use disorders. This study was approved as an institutional review board (IRB) exemption by the local VA institutional review board (IRB 1223219).

### Treatments

Ketamine was administered via either the intranasal (esketamine, n=99), intravenous (racemic, n=20), or intramuscular (racemic, n=1) routes. Individual patients received the same route and formulation of ketamine for all sessions. Intranasal esketamine doses typically started at 56 mg and were increased to 84 mg after the initial session. Intravenous doses had more variability, as they were dosed at 0.5 to 1 mg/kg, with doses adjusted based on tolerability, side effects and efficacy. Formal psychotherapy was not paired with ketamine sessions, though psychological support was available if needed. Most patients (110 of 120) completed at least eight treatment sessions, with the remaining completing between 2 and 7 sessions each.

### Analysis of average PHQ-9 scores

Patients completed PHQ-9 questionnaires at baseline and prior to ketamine treatment sessions. Supplemental Table 1 lists the full text for the nine items of the PHQ-9. Average PHQ-9 sum scores across patients were computed across the first eight treatment sessions. Separately, average PHQ-9 item responses were computed across the eight sessions. Significant change in sum score was defined as p < 0.05 on the Friedman chi-square test for repeated measures, as the first two sessions’ data violated normality (p < 0.05, Shapiro-Wilk test). Significant change in item response was defined as p < 0.006 (after Bonferroni correction for multiple comparisons) on the Friedman test. Only patients who completed a PHQ-9 before all eight sessions (82 of 120) were included in these repeated measures tests.

### Analysis of item trajectories

Item responses were analyzed across treatment sessions for each item, *i*, and for each patient, *p*. Linear (Equation 1) and exponential (Equation 2) models were fit to item response trajectories by minimizing the residual sum-squared errors (RSS). In the linear model, *m* is the linear slope, *t* is time in days, and *b* is the intercept. In the exponential model, *a* is a scaling factor, *m* is a growth factor, *t* is time in days, and *b* is a constant offset. In both models, *m* reflects a rate of change in item response.

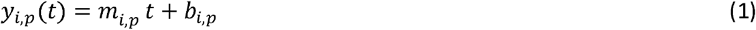

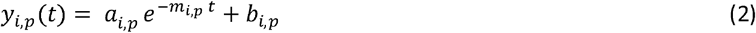

For each model, the Akaike information criterion (AIC) and Bayesian information criterion (BIC) were computed for the purposes of model selection. Specifically, AIC and BIC were computed using the RSS from model fitting, assuming independent and identically distributed (IID) residuals following a normal distribution. For each patient, the winning model was defined as the model with lowest AIC and BIC for the majority of items. In turn, the best overall model for a given questionnaire was defined as the winning model for the majority of patients. The linear model, as the best overall, was used as the basis of all subsequent analyses. Differences in linear slopes between the items was defined as p < 0.05 on the Friedman test for repeated measures, with post-hoc evaluation for pairwise differences via Wilcoxon signed-rank test with Bonferroni correction for all pairwise comparisons (p < 0.05 / 36 = 0.0014). Effect sizes for pairwise differences was defined as the difference in medians divided by the average of the two interquartile range (IQR) values [26].

### Principal component analysis for linear slopes

Using linear models (Equation 1), each patient had nine slope parameters describing their change in each of the PHQ-9 items. Principal components analysis (PCA) was computed in the 9-dimensional space of slopes. PCA finds the set of orthogonal linear combinations which capture maximal variance across participants. Parallel analysis was used to evaluate the strength of principal components (PCs), in which the actual eigenvalue of each PC was compared with the 95^th^ percentile of the distribution of eigenvalues for that PC from 10,000 randomly generated datasets. Coefficients for each significant component were analyzed to understand directions of change.

### Predicting treatment response

Treatment response was defined via the sign for the first principal component (PC1), which by definition captures the majority of variance in the data, and in our case represented a weighted rate of change in symptoms. Due to data normalization, a negative sign implies a greater improvement along PC1 than the mean across participants. To relate this to classical measures of response, percent changes in PHQ-9 sum scores were also evaluated. Logistic regression was used to predict treatment response using patients’ baseline PHQ-9 item responses. An exhaustive feature selection was conducted to evaluate all 511 possible subsets of PHQ-9 items as features. For each model, classification performance was evaluated across 1000 iterations of repeated 5-fold cross validation.

### Threshold tuning for high confidence predictions

Logistic regression provides probabilities of test items belonging to each class. The choice of classification threshold results in a tradeoff between positive predictive value (PPV) and sensitivity, or between negative predictive value (NPV) and specificity. For all 511 possible subsets of features, a parametric search was conducted across 20 different classification thresholds ranging from 0 to 1. This resulted in over 10,000 distinct classification models, which then underwent cross validation as before. To optimize for potential clinical utility, models were selected that met a performance specification of either: (1) at least 90% PPV and at least 10% sensitivity for predicting “response”, or (2) at least 90% NPV and at least 10% specificity for predicting “non-response”. These criteria were chosen because very high predictive value could provide clinically actionable information, even if only for a subset of the patients. In contrast, models with lower predictive value are unlikely to change clinical management, even if they have higher sensitivity. Among the models meeting these specifications, performance was ranked by the arithmetic mean of predictive value (PPV or NPV) and coverage of the relevant cases (sensitivity or specificity), with the best model defined as the one with highest mean of those two characteristics.

### Code and data availability

All data analysis was conducted with custom Python software utilizing open-source scientific and machine learning packages. The code to reproduce all results and figures is available at github.com/angevineMiller/ketaminePrediction. Data are available upon request.

## Results

This study was motivated by two major goals. First, we sought to understand the dynamics of depression symptoms over the course of repeated ketamine sessions. Second, using a symptom-level modeling approach, we aimed to predict treatment response for patients in a real-world clinical setting. We first confirmed that average PHQ-9 sum scores improved across the ketamine treatment sessions (Figure 1a) (p < 10^-18^, Friedman test). In addition, average responses for every individual PHQ-9 item improved over the course of treatment (Figure 1b) (ps < 10^-5^, Friedman test). We then proceeded to analyze symptom trajectories for each individual patient.

**Figure 1.**
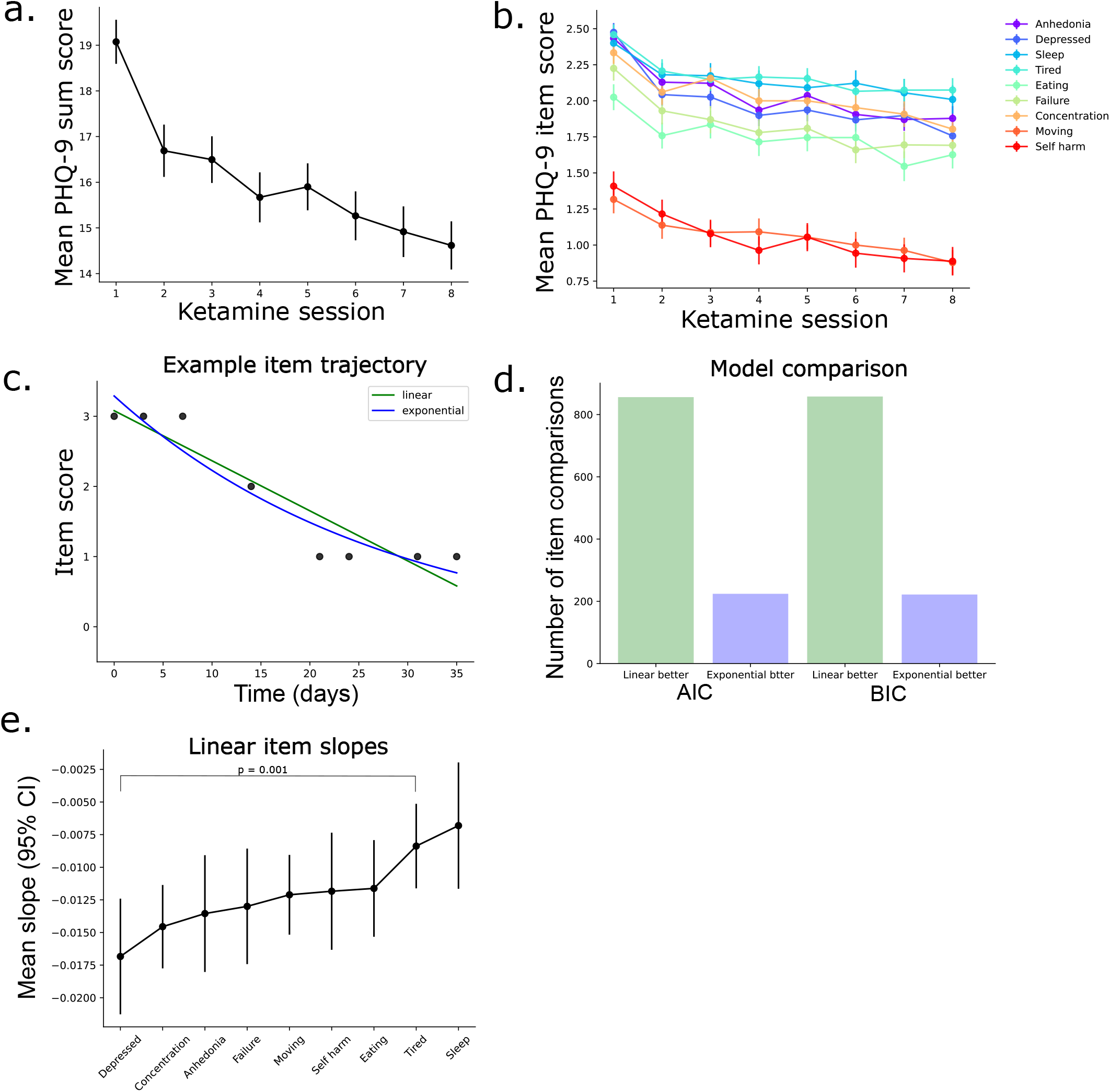
Effects of ketamine on individual symptoms of depression. (a) Mean (SEM) PHQ-9 sum scores across patients at each of the first eight ketamine sessions. (b) Mean (SEM) PHQ-9 item scores across patients at each of the first eight ketamine sessions. (c) One example patient’s item responses for one example item (PHQ-9 item #1). Green and blue curves are the linear and exponential model fits, respectively. (See Methods for model equations). (d) Bar plots comparing linear and exponential model fits using AIC (left) and BIC (right), such that the bar heights indicate the number of patient-items where the linear (green) or exponential (blue) model was a better fit. (e) Mean (95% CI) linear slopes across patients for each of the PHQ-9 items. P-value shown for pairwise comparison with significant difference after Bonferroni correction for multiple comparisons. See Supplemental Table 1 for mapping of item names to PHQ-9 item text.

### Modeling individual symptom trajectories across individuals

We sought the simplest model that could capture symptom changes over the course of treatment in the majority of individuals. In addition to a simple linear model (Equation 1), we also tested an exponential model (Equation 2), motivated by prior work showing that exponential functions can describe various interventions for depression [27,28]. We fit linear and exponential models to item response trajectories for each patient (Figure 1c), and then evaluated the fit of both models using AIC and BIC. Linear models were better than exponential models for the majority of patients (104 of 120 for AIC, 105 of 120 for BIC) (Figure 1d), so we chose the linear model as the basis for further analysis to maintain consistency/comparability across individuals.

The slopes of the linear models describe the rate of change in each symptom of depression for patients over the course of their ketamine treatment. The average slopes across patients were negative for all nine items, confirming that patients generally improved across all symptoms (Figure 1e). We detected a difference in slopes between the items (p = 0.012, Friedman test). Post-hoc comparisons revealed that PHQ-9 item #2 (depressed mood) had significantly steeper slopes than item #4 (tiredness) after Bonferroni correction (p = 0.001, Wilcoxon signed-rank test) (Figure 1e). Thus, symptoms of depressed mood improved more rapidly than tiredness across ketamine treatment, with a difference in medians of about 45% of the interquartile range (see Methods).

### Low Dimensional Variables to Capture Trajectory Changes

Next, we wondered whether we could parsimoniously capture how patients varied in the high-dimensional space of symptom trajectories. To understand this we computed a principal components analysis (PCA) on the trajectory slopes calculated for each item of the PHQ-9 across subjects. We detected two significant principal components (PCs), which explained 52.2% and 16.2% of the variance, respectively (significance calculated using parallel analysis, see Methods) (Figure 2a). Analysis of coefficients revealed that the first PC (PC1) described a weighted rate of change for all nine symptoms in the same direction (Figure 2b). Coefficients of the second PC (PC2) showed opposite signs for somatic symptoms (e.g. appetite, energy, concentration, movement) and affective symptoms (e.g. mood, anhedonia, thoughts of self-harm or of being a failure) (Figure 2c). Projecting participants’ slopes data onto these two PCs revealed the distribution of patients across these two dimensions of symptom variance (Figure 2d).

**Figure 2.**
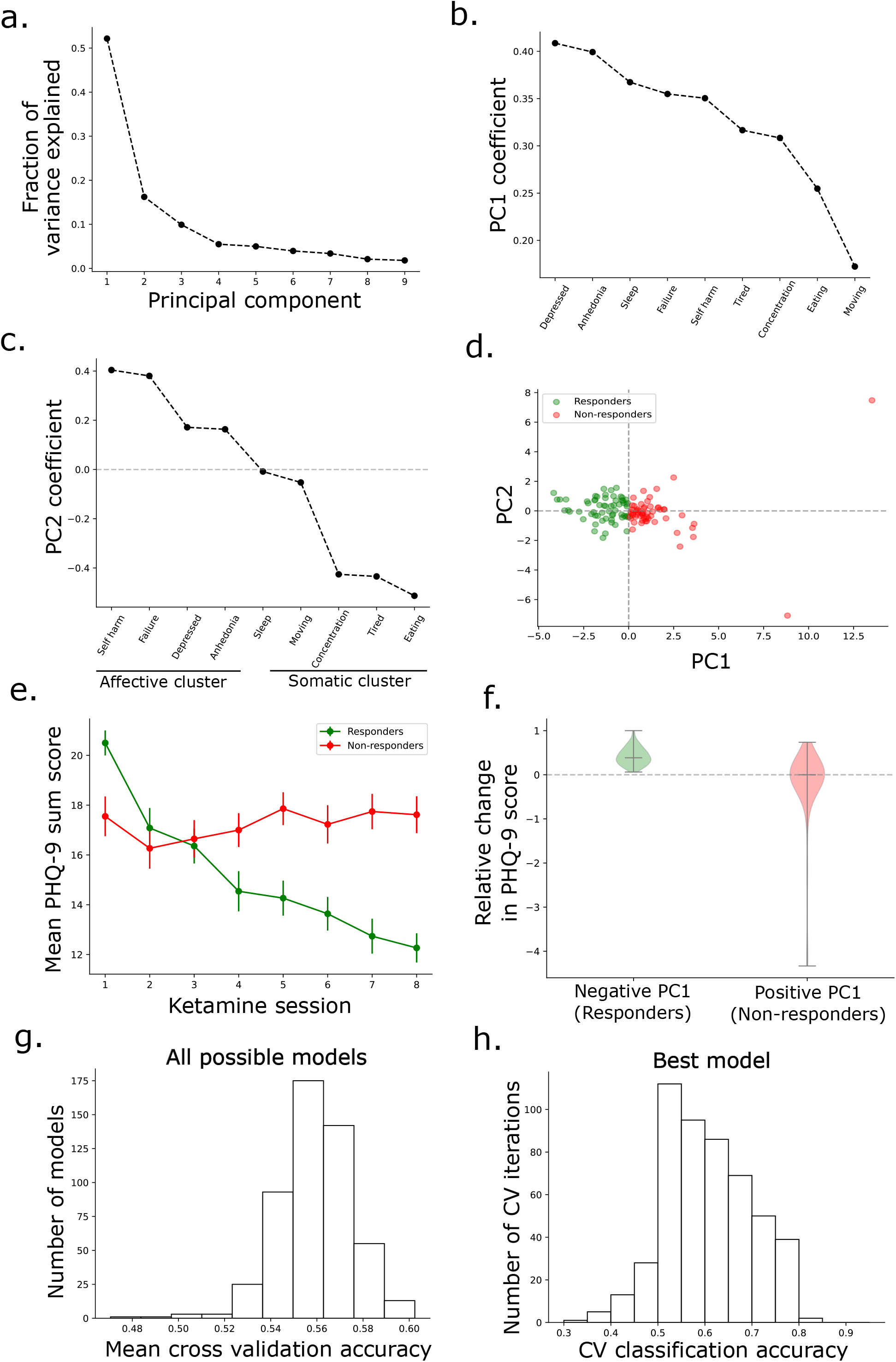
Predicting treatment response from baseline symptoms. (a) Scree plot showing fraction of total variance explained by each principal component (PC) from a PCA over the nine-dimensional data of linear slopes for each patient. Significance of PCs was determined by bootstrapping with parallel analysis (see Methods). (b) Coefficients of the first PC of the data for each of the features, corresponding to PHQ-9 item slopes. (c) Coefficients of the second PC of the data for each of the features, corresponding to PHQ-9 item slopes. (d) Scatter plot of the projections of each patient’s set of linear slopes onto the first two principal components of the data, colored according to the sign of the first principal component. Responders (green) are defined as having a negative sign of the first PC, while non-responders (red) are defined as having a positive sign of the first PC. (e) Mean (SEM) PHQ-9 sum scores across patients for the first eight ketamine sessions for responders and non-responders. (f) Violin plots of relative change in PHQ-9 sum score from baseline to last session for responders (green) and non-responders (red). Bars indicate median and extreme values, and width of violin indicates distribution density. Dashed horizontal line indicates 0 change. (g) Histogram of the mean cross-validation accuracy for all models from an exhaustive feature selection over all possible subsets of baseline PHQ-9 items as model features. (h) Histogram of accuracies across all iterations of cross-validation for the best model from the exhaustive feature selection, i.e. the model with overall best mean CV accuracy. Inset lists the other key model performance characteristics for this model. For (b) and (c), see Supplemental Table 1 for mapping of item names to PHQ-9 item text.

Based on the coefficient weights, the sign of the first principal component (PC1) represented whether a patient improved more (negative sign) or less (positive sign) than the mean rate of change in symptoms (Figure 2d). This is because each patient’s symptom slopes were standardized by the population mean slopes prior to computing PCA. Therefore, this offers a simple, data-driven way of differentiating treatment responders from non-responders. To better illustrate this, we first plotted average PHQ-9 sum scores for patients with negative PC1 (identified as responders, n=62) compared to patients with non-negative PC1 (identified here as non-responders, n=58) (Figure 2e). The sum scores among responders improved significantly between ketamine induction sessions (F(7,336) = 46.7, p < 10^-45^, partial eta-squared = 0.49, repeated measures ANOVA). The mean change in sum scores between baseline and the final session among responders was -8.7 points (95% CI: -9.8 to -7.6). In contrast, we detected no differences in sum scores between sessions among the non-responders (F(7,224) = 2.07, p = 0.07, Greenhouse-Geisser correction, repeated measures ANOVA). The mean change in sum scores among non-responders was -0.40 (95% CI: -2.0 to 1.2).

Based on the distribution of PC2 coefficients across symptom subdomains, we hypothesized that the sign of the 2^nd^ PC (PC2) could reflect whether there were relatively greater improvements in the affective symptom clusters (negative sign) or somatic symptom clusters (non-negative sign). To test this, we calculated (for responders only) the trajectories of average scores for the affective and somatic subdomains of symptoms, grouped using the PC2 weights noted above. As expected, responders with a non-negative PC2 showed greater improvements in somatic symptoms, compared to responders with a negative PC2 (F(7, 329) = 3.03, p = 0.004, mixed ANOVA interaction) (Supplemental Figure 1a). In contrast, we were unable to detect a significant difference in the rate of improvement in affective symptoms based on PC2 sign (F(7, 329) = 1.01, p = 0.42, mixed ANOVA interaction) (Supplemental Figure 1b). Thus, the sign of PC2 reliably differentiated patients based on their rate of improvement on the somatic subdomain, suggesting improvement in this domain of symptoms reflected another meaningful source of variance even within those who were categorized as responding overall.

### Predicting response from baseline symptoms

We identified above a data-driven approach to characterizing the antidepressant response to ketamine treatments using changes in individual item scores of the PHQ-9. Traditional measures of treatment response are defined with respect to the percent change in the overall PHQ-9 sum score. To ensure that our categorization corresponds at least roughly with more standard definitions of treatment response, we computed the percent reduction in PHQ-9 sum score from baseline to the last treatment session for patients categorized by negative PC1 (responders) and non-negative PC1 (non-responders). Among responders, the median percent change in PHQ-9 sum scores was a reduction by 39% (IQR 25%). Among non-responders, the median percent change in sum scores was 0% (IQR 22%) (Figure 2f), suggesting that in general this method of classifying subjects was reasonable.

Thus, using this categorization, we next wanted to see if baseline symptoms might provide some ability to classify responders and non-responders. Prior to classification, we first analyzed whether PC1 scores were related to basic demographics (age, gender) and treatment type (IV racemic ketamine vs. intranasal esketamine). We detected no differences in average PC1 value based on ketamine formulation (esketamine vs IV racemic) (p = 0.59, two-sided t-test), gender (p = 0.19, two-sided t-test) or age (r = 0.13, p = 0.17, Pearson’s correlation), suggesting these simple demographic factors would not be informative for group membership or classification (data not shown). We next asked whether treatment response, as defined above using change in PC1, could be predicted from single item PHQ-9 scores alone. Specifically, we used logistic regression to predict the sign of PC1 using baseline PHQ-9 items as features. To identify the best features for prediction, we conducted an exhaustive feature selection by fitting logistic regressions with all 511 possible subsets of PHQ-9 items as features. For each of these 511 models, performance was evaluated with repeated 5-fold cross-validation (CV).

Nearly all of the models (509 of 511, 99.6%) had better-than-chance average CV accuracy, though all were only moderately better than chance (Figure 2g). The model with best CV accuracy used only two PHQ-9 items as features: item #1 (anhedonia) and item #6 (feeling of failure). Higher baseline responses on either item predicted subsequent treatment response. For this model, the average classification accuracy to holdout data was 60.3% (95% confidence interval 59.5 – 61.1%) (Figure 2h). The precision (PPV) was 60.2%, recall (sensitivity) was 68.3%, and F1 score was 64.0%. The second-best model used only one item as a feature, item #2 (depressed mood), with nearly as good performance as the best model: 60.0% average accuracy (95% CI 59.2% -60.8%), 60.0% precision, 67.7% recall, and 63.6% F1 score. Patients with higher baseline depressed mood were more likely to respond.

### Threshold tuning for high confidence predictions

Although the above approach identified many models that can predict treatment response better than chance, none of those models had excellent performance characteristics. They had a maximum of around 60% predictive value, which is unlikely to provide sufficient prediction confidence for changing clinical management. We reasoned that a more clinically useful model would have a very high predictive value (PPV or NPV). Such a model would provide clinically actionable information, even if only for a subset of the patients. For example, if a model could predict with over 90% NPV that a patient will not respond to ketamine, then we could be quite confident in recommending an alternative treatment for that patient.

Seeking such a model, we used threshold tuning in order to optimize the classifiers toward high predictive confidence (Figure 3a). Specifically, instead of using probability 0.5 as the threshold to classify patients as responders and non-responders, we evaluated a range of thresholds between 0 to 1 (see Methods). Changing the threshold in this way necessarily results in a tradeoff between prediction confidence (PPV or NPV) and coverage of the relevant cases (sensitivity or specificity). For model selection, we conducted another exhaustive search across all possible subsets of baseline PHQ-9 items, and for each of these feature sets, we applied threshold tuning to evaluate a range of thresholds (Figure 3b). This resulted in over 10,000 distinct models that were cross validated to evaluate model performance on holdout data (Figure 3c).

**Figure 3.**
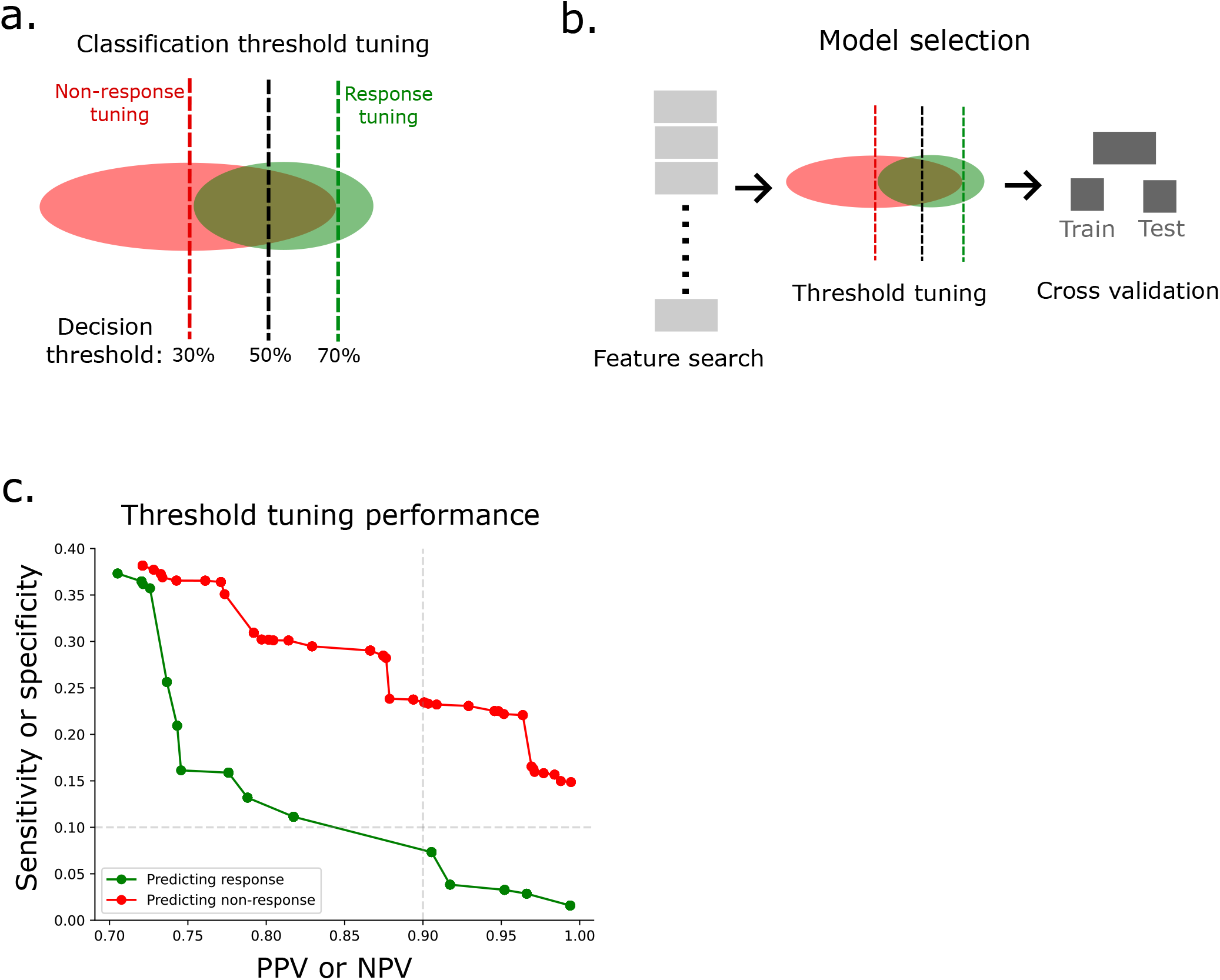
Threshold tuning to confidently predict non-response. (a) Diagram depicting threshold tuning approach. Red and green ellipses represent a hypothetical projection of data for non-responders and responders, respectively. Black dashed line indicates standard decision threshold for logistic regression, which maximizes accuracy. Green and red dashed lines indicate alternate choices for the decision threshold, which maximize positive predictive value (PPV) for predicting response or negative predictive value (NPV) for predicting non-response, respectively. (b) Diagram depicting three major steps of model selection procedure: feature search, threshold tuning, and cross validation. Grey boxes represent the different sets of baseline PHQ-9 items that can be tried as a feature set. Threshold tuning, as in (a), was conducted for each feature sets, resulting in over 10,000 models that then underwent cross validation. (c) Scatter plot of cross validation model performance for some of the best models for predicting response (green) or non-response (red). Plot shows the inherent tradeoff between predictive value (PPV or NPV) and coverage of the relevant cases (sensitivity or specificity). Green dots represent the best models, in terms of highest sensitivity (y-axis), among all of the models with at least a minimum PPV (x-axis). Red dots represent the best models, in terms of highest specificity (y-axis), among all of the models with at least a minimum NPV (x-axis). Dashed grey lines show cutoffs for our predefined model performance specifications: PPV or NPV > 90%, sensitivity or specificity > 10% (upper right quadrant).

We then selected only those models with very high predictive value, while at the same time retaining a minimum coverage of the relevant cases (Figure 3b, upper right quadrant). Specifically, we searched for models with either (1) at least 90% PPV and at least 10% sensitivity, or (2) at least 90% NPV and at least 10% specificity. We identified hundreds of models meeting these performance criteria. The best of these models (see Methods) had a NPV of 96.4%, while retaining a specificity of 22.1%. This model used only three baseline PHQ-9 items as features: item #2 (depressed mood), item #5 (changes in appetite or eating), and item #9 (self-harm or suicidal ideation). Relatively speaking, lower values on item #2 and item #5 and higher values on item #9 favored non-response (see Supplemental Methods).

Thus, using threshold tuning, we were able to identify classification models that predict non-response with very high confidence (over 96% NPV), based purely on patients’ baseline PHQ-9 symptom scores. In contrast, we detected no models for predicting treatment response at the specified performance cutoffs (Figure 3c, green).

## Discussion

In this study we developed an approach for modeling changes in symptoms of depression over the course of repeated ketamine sessions. Using this approach, we found that all symptoms improved across the course of ketamine treatment, and the symptom of depressed mood improved more rapidly than the symptom of low energy (Figure 1). We found a range of individual differences in these item response trajectories, both in the degree of change in overall depression level, and in specific subdomains of symptoms (Figure 2). We developed logistic regression classifiers, which can predict better than chance whether patients will respond to ketamine, using their baseline symptoms alone (Figure 2). Finally, using threshold tuning, we found classifiers that can identify a subset of patients who are highly *unlikely* to respond to ketamine with over 96% predictive value (Figure 3). Our findings shed light on how ketamine affects specific symptoms and dimensional features of depression, and the method for identifying non-responders may prove useful for informing rational treatment recommendations among a growing list of novel therapeutics for treatment resistant depression [11–14].

Few prior studies have analyzed how different depression symptoms respond to ketamine, particularly across repeated doses. Floden and colleagues (2022) [21] conducted a symptom-level analysis of data from the TRANSFORM 2 trial of repeated intranasal esketamine for TRD. They found that eight twice-weekly doses of esketamine plus oral antidepressant led to improvements in all PHQ-9 items except, interestingly, not item #9 concerning suicidality [21]. They suggest that this could be due to low levels of baseline suicidality due to exclusion of patients at serious risk of suicide from the TRANSFORM 2 trial. In contrast, our data show clear reductions in suicidality, highlighting the value of studying outcomes in heterogeneous real-world data with complex patients.

We found that all depression symptoms improved across repeated ketamine sessions, but the symptom of depressed mood improved faster than the symptom of low energy. This supports results from Chen et al. (2021) [23], who found that a single infusion of low-dose IV ketamine resulted in greater reductions in cognitive and affective symptoms, compared to somatic symptoms [23]. Similarly, Park and colleagues (2020) [22] found that typical/melancholic symptoms improved more rapidly than atypical symptoms of depression after a single dose of IV ketamine [22]. Furthermore, using principal component analysis (PCA), we found significant individual variation in whether patients improved more in affective symptoms (e.g. depressed mood, anhedonia, suicidal ideation) or in somatic symptoms (e.g. changes in appetite, concentration, or energy level) suggesting that symptom response to ketamine is patient-specific.

Using logistic regression classifiers, we were able to predict which patients would respond better than chance using their baseline symptoms alone. This was likely possible due to a difference in baseline depression severity, such that responders had more severe baseline depression than non-responders. These results may be counterintuitive, as one might expect patients with more severe depression to be more treatment resistant. A study by Jesus-Nunes et al. (2022) [29] found that patients with more severe depression were less likely to respond to a single infusion of IV ketamine or esketamine. On the other hand, our data may reflect the consistent finding that antidepressants separate from placebo most prominently among patients with severe baseline depression [30–32], which has also been observed for ketamine [33]. It is unclear how much of that is due to stronger effects of the active drug or weaker effects of the placebo among patients with more severe depression. It is also possible that baseline depression severity may signal different underlying psychopathologies, with consequent differences in response to ketamine, but future studies will be necessary to explore that possibility.

Importantly, the standard classifiers noted above are not likely to be useful in the clinic as they can only predict response with about 60% positive predictive value. We reasoned that a much higher predictive value would be required for a model to guide clinical decision making. We therefore optimized the classifiers for either high PPV for identifying responders, or for high NPV for identifying non-responders. We did this via threshold tuning, in which we searched over the full space of decision thresholds, and then selected classifiers with optimal cross-validated performance characteristics. Using this approach, we found models that could predict *non-response* to ketamine, i.e. treatment failure, with close to perfect negative predictive value. Although this comes at the expense of a reduced specificity, the model could prove useful because it provides directly actionable information for those patients that it identifies as non-responders. Future pre-registered and controlled studies will be necessary to test whether these findings replicate and generalize. This simple approach of threshold tuning to find clinically meaningful predictions may be useful for other treatments as well.

## Limitations

There are several important limitations of this study. First, this is a non-registered secondary analysis, so the results are fundamentally exploratory and require confirmation with pre-registered and experimental studies. These are real world clinical data without placebo or wait list control groups, making it impossible to distinguish the role of nonspecific factors including placebo. For the same reason, we cannot exclude the role of regression to the mean, which is a possible explanation for observing improvement among patients with more severe baseline depression. However, regression to the mean is unlikely to be the only driver of this finding, since the average trajectories for responders and non-responders did not converge to a common mean value. It is also important to highlight that psychotherapy was not provided in concert with ketamine treatments, so our data do not speak to the potential for ketamine-assisted psychotherapy to prolonging or change its effects [34]. Finally, we lack detailed patient accounts of their treatment, which would be helpful for understanding patient-centered and functional outcomes beyond those limited data that are reflected on the PHQ-9 questionnaire.

## Conclusion

Repeated ketamine and esketamine led to progressive improvements in all symptoms of depression, but depressed mood improved faster than low energy. Using machine learning classifiers, we could predict non-response to ketamine with very high confidence for a subset of the patients. If validated in future studies, this model could be useful for identifying patients unlikely to benefit from ketamine, who might be better served by other treatments.

## Data Availability

All data produced in the present study are available upon reasonable request to the authors

## Acknowledgment

D.R. is supported by funding from the Center of Excellence for Stress and Mental Health, Burroughs Wellcome Fund, Career Award for Medical Scientists.

