## Supplemental Materials for "Predicting non-response to ketamine for depression: a symptom-level analysis of real-world data"

**Supplemental Information**

**Supplemental Table 1. PHQ-9 items**

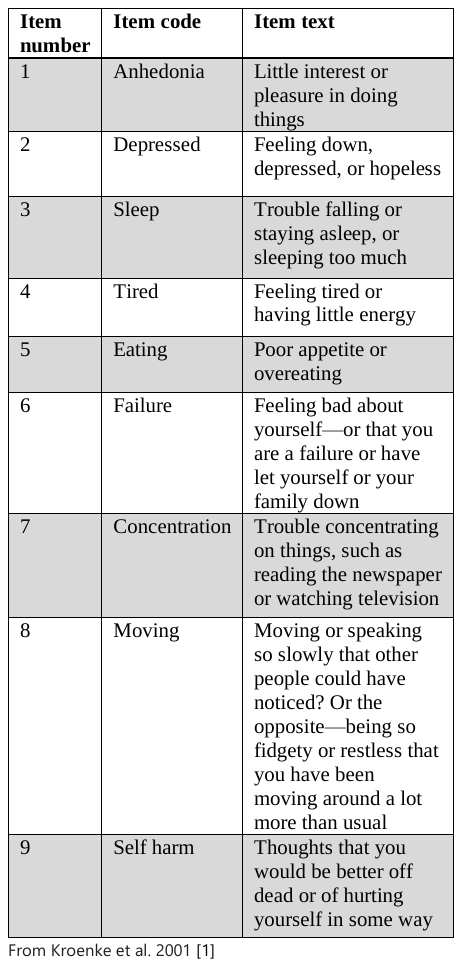

**Supplemental Figure 1. Somatic and affective subdomain trajectories**

(a) Mean (SEM) sum scores for somatic subdomain symptoms across only the responders (i.e. only patients with positive sign PC1) across treatment sessions, grouped by those with negative PC2 sign (dashed black line) and those with non-negative PC2 sign (solid black line). (b) Mean (SEM) sum scores for affective subdomain symptoms across only the responders, grouped by those with negative PC2 sign (dashed black line) and those with non-negative PC2 sign (solid black line).

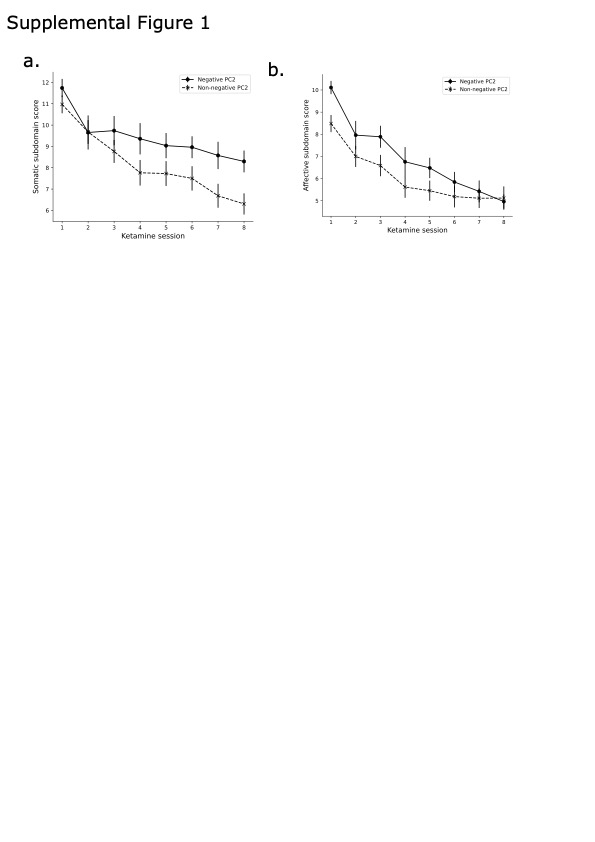

**Supplemental Methods**

*Relatively more sleep trouble and suicidal ideation among non-responders*

Analyzing the coefficients of the best model for predicting non-response, we noticed that one of the features (item #9, self-harm and suicidal ideation) had a negative regression coefficient. This implies that higher baseline values on this feature predict non-response, the opposite of the other two features. To determine the generality of this observation, we plotted distributions of coefficients across all of the models with >90% NPV and >10% specificity. Item #9 had negative coefficients for all of these models, as did item #2 which concerns sleep difficulties. In contrast, the other seven items had positive coefficients across the vast majority of models. In other words, higher baseline scores on items #2 and #9, relatively to the other items, predicts non-response. However, importantly, these results do not imply that non-responders have higher baseline scores on any of these items. In fact, patients classified as non-responders have lower average baseline responses on all PHQ-9 items, including item #2 and item #9. Rather, this finding suggests that *relatively speaking* non-responders’ symptoms are weighted more towards sleep difficulties and suicidal ideation compared to the other symptoms. This finding is consistent, but far less salient than the more obvious finding of higher baseline depression severity among the responders.
